# Wearable technologies for children with chronic illness: A Proof-of-Concept Study

**DOI:** 10.1101/2020.10.25.20219139

**Authors:** Flora McErlane, Elin Haf Davies, Cecile Ollivier, Anna Mayhew, Obuchinezia Anyanwu, Victoria Harbottle, Aimee Donald

## Abstract

**Objective:** To determine the feasibility of wearable technologies in physical activity assessment in three paediatric diseases, namely Niemann-Pick C (NP-C), Juvenile Idiopathic Arthritis (JIA) and Duchenne Muscular Dystrophy (DMD).

**Design:** Proof of concept feasibility study

**Setting and patients:** Thirty children were recruited across three UK hospitals (Royal Manchester’s Children Hospital, Great Ormond Street Children’s Hospital and the Great North Children’s Hospital). Ten were diagnosed with NP-C, eight with DMD and twelve with JIA.

**Intervention:** All participants completed the 6-minute walk test (6MWT) at enrolment. Patients were provided with disease specific smartphone apps paired with a wearable device via Bluetooth. Ambulation was recorded in 30-minute epochs measuring average daily maximum (ADM), average daily steps (ADS), and average daily steps per 30-minute epoch (ASE).

**Results:** Median 6MWT results were 450m, 325m and 434.5m for the NP-C, DMD and JIA cohorts respectively. Wearable data capture was feasible in all three disease groups, although complete data capture was not sustained. A statistically significant between-cohort difference was identified for ADM, ADS and ASE. Statistically significant differences were found between DMD/JIA for ADM; NP-C/DMD for ADS and DMD/JIA for ASE.

**Discussion:** Wearable sensor technologies have the potential to add important information to our understanding of ambulation in chronic paediatric disease. The wearable devices were easy to use and popular with patients although key features need to be addressed to ensure higher engagement in future deployments. As the technology continues to evolve at a rapid pace, opportunities to implement child friendly solutions are already available.

## Introduction

Recent advances in modern technologies based on wearables, sensors or videos allow the acquisition of continuous real-time datasets relating to many aspects of an individual’s life. Wearable technologies such as accelerometers may be able to complement traditional clinical outcome assessments (COA) in paediatric research, addressing both clinical and research needs in a patient-centred and minimally intrusive fashion.

Clinicians and clinical researchers are increasingly interested in the potential impact of such data streams on patient care and service delivery [1]. Wearable technologies have been used to quantify activity levels in healthy children [2] as well as in many different conditions [3]. The use of innovative digital tools in clinical research is supported by FDA [4] [5] and EMA [6].

With this in mind, we hypothesised that wearable technologies may be a valid alternative to current physical activity measurement techniques or surrogate measures of activity such as the 6-minute walk test (6MWT) for measuring ambulatory activity in children and young people (CYP). We present a proof of concept evaluation to determine the feasibility of using a wearable device to assess physical activity in three different paediatric diseases, namely Niemann-Pick C (NP-C), Juvenile Idiopathic Arthritis (JIA) and Duchenne Muscular Dystrophy (DMD). While the three diseases have different pathologies (neurological, musculoskeletal and neuromuscular), each may have a significant impact on gait and ambulatory ability.

Ethical approval was granted by the NRES Committee London - City & East and Bristol Research Ethics Committee Centre (REC reference: 15/LO/0528; IRAS project ID: 174833). Patients were recruited from Royal Manchester’s Children Hospital, Great Ormond Street Children’s Hospital and the Great North Children’s Hospital in the UK. Parental consent was obtained, and child assent where relevant.

### Juvenile Idiopathic Arthritis

Juvenile Idiopathic Arthritis (JIA) is the most common rheumatological diagnosis in childhood, with a prevalence of approximately 1 in 1000 children in the UK [7]. Many CYP will experience arthritis in their lower limbs during the disease course, with the knee and ankle being common sites of synovitis [8]. Studies of gait and mobility in CYP with JIA suggest that some individuals exhibit abnormal gait patterns secondary to their disease, with this having a subsequent effect on their day-to-day activities [9].

CYP with JIA may be less active than their healthy peers with an inverse relationship between physical activity and pain [10-11]. A recent study gathering the perspectives of CYP with JIA and their parents relating to physical activity highlights pain as the most important barrier to physical activity, and concludes that CYP should be taught strategies to manage their pain whilst maintaining physical activity levels [11].

### Niemann-Pick C disease

Niemann-Pick type C disease is a rare, neurodegenerative, autosomal recessive disorder that primarily manifests in children. NP-C patients who survive infancy experience progressive neurodegeneration of the brain, manifesting as cerebellar ataxia (present in 70% of patients from the International NPC Registry) [12]. This is symptomatically expressed as a disturbance of gait, balance and coordination resulting in recurrent falls, clumsiness and limb ataxia, [13]. As the disease progresses ataxia symptoms become more disabling, severely impairing quality of life and functioning, and leading to a loss of ambulation and an increase of the utilization of health resources [14]. The work was a collaborative effort with two patient groups (Niemann Pick Association UK and International Niemann Pick Disease Alliance (INPDA)) and clinicians.

### Duchenne Muscular Dystrophy

DMD is the most common childhood neuromuscular disorder, affecting 1 in 5,000 boys [15]. It is an X-linked disorder caused by mutations in the dystrophin gene and impacts skeletal, respiratory and cardiac muscle. This progressive muscle weakness is usually identified in the early toddler years when the child experiences motor difficulties and may also present with delayed motor milestones [16]. With improved standards of care and the use of glucocorticoids, loss of ambulation occurs generally around the age of 12 - 14 [17], and survival is up to the third and fourth decade. This pilot study was funded and designed in collaboration with Duchenne UK, a patient advocacy group.

## Methods

Patients were identified and recruited by their UK clinical care teams at Manchester’s Children Hospital, Great Ormond Street Children’s Hospital and Newcastle Hospitals Foundation Trust. Aparito configured a study-specific solution and provided a paired app and wearable device for each disease, capturing real-time data and stored data on the band’s memory until transfer by Bluetooth to a secure database.

Preliminary demographic data, motor developmental data and current activity levels were collected at baseline. All participants completed the 6MWT at enrolment, as per American Thoracic Society guidelines [18]. 6MWT results were compared with age-appropriate predicted results obtained from healthy children in Switzerland [19].

Each patient was allocated a wrist-worn wearable device (3D accelerometer) that records physical activity (measured in steps and distance) paired with an application downloaded to the parents/carer mobile phone/tablet (Android and iOS) and directly on to the young person’s own phone in some cases. Accelerometer data were visible to the research team but not the CYP or their family. Participants’ step counts were compared with age-appropriate recommended daily step counts [20].

In the DMD and JIA studies, CYP were asked to wear the device continuously for 12 weeks but the NP-C study patients were asked to wear the device with no predefined set duration. The wrist worn wearable device was the same in each study and captured data in 30 minutes epochs. The 30 minutes epochs were used to provide a three-month battery life, and reduce the burden of charging. Sensor data were analysed in three different domains

1. Average Daily Maximum (ADM): The mean of daily epochs with the most steps, calculated over a month
2. Average Daily Steps (ADS): The mean of the total number of daily steps, calculated over a month
3. Average Steps per Epoch (ASE): The mean number of steps in each 30-min epoch, calculated over a day. Active days (days with >4 hours of data) were then averaged over a month.

As part of this feasibility study, clinic-based (6MWT) and wearable data were analysed for group differences between disease cohorts. A non-parametric assessment of differences between the three disease groups was conducted for the 6MWT; the ADM, ADS and ASE were compared between disease groups (Kruskal-Wallis test) and explored with Dunn’s multiple comparison test. Spearman’s rank correlation coefficient was applied to identify any association between in-clinic 6MWT and wearable metrics.

## Results

### Demographic information and descriptive statistics

Across all three studies a total of 30 CYP were recruited, 10 diagnosed with NP-C, 8 with DMD and 12 with JIA. Twenty used the wearable to share data. Age ranged from 6-16 years, with the NP-C children having a younger median age of 10 years of age, and the JIA cohort the oldest with a median age of 13.5 years. Table 1 shows demographics, 6MWT and wearable metrics.

**Table 1:**
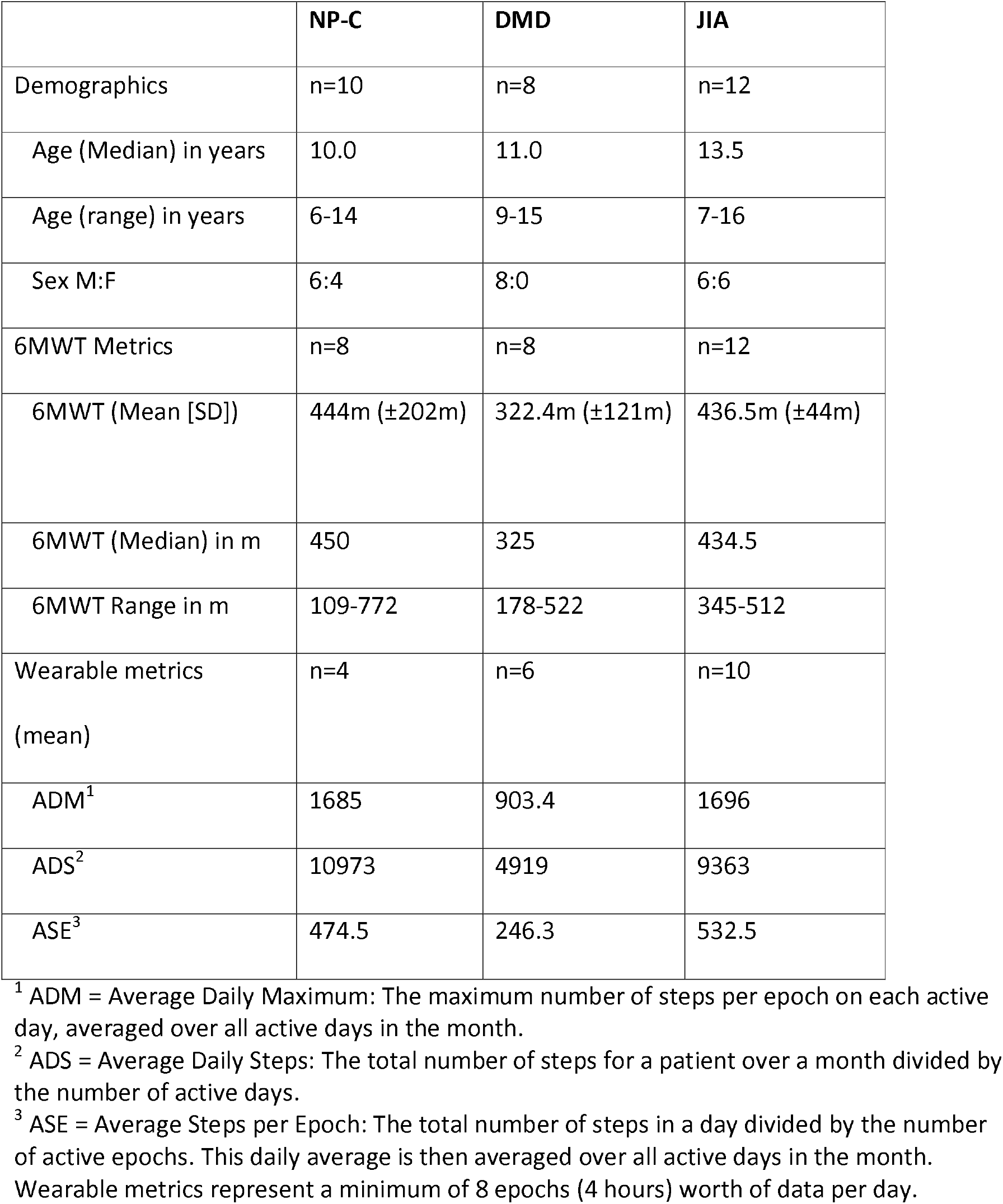
Demographics, 6MWT and wearable metrics for month 0-1 from patients enrolled in the studies.

|  | **NP-C** | **DMD** | **JIA** |
| --- | --- | --- | --- |
| Demographics | n=10 | n=8 | n=12 |
| Age (Median) in years | 10.0 | 11.0 | 13.5 |
| Age (range) in years | 6-14 | 9-15 | 7-16 |
| Sex M:F | 6:4 | 8:0 | 6:6 |
| 6MWT Metrics | n=8 | n=8 | n=12 |
| 6MWT (Mean [SD]) | 444m ($\pm 202$m) | 322.4m ($\pm 121$m) | 436.5m ($\pm 44$m) |
| 6MWT (Median) in m | 450 | 325 | 434.5 |
| 6MWT Range in m | 109-772 | 178-522 | 345-512 |
| Wearable metrics<br>(mean) | n=4 | n=6 | n=10 |
| ADM<sup>1</sup> | 1685 | 903.4 | 1696 |
| ADS<sup>2</sup> | 10973 | 4919 | 9363 |
| ASE<sup>3</sup> | 474.5 | 246.3 | 532.5 |

<sup>1</sup> ADM = Average Daily Maximum: The maximum number of steps per epoch on each active day, averaged over all active days in the month.
<sup>2</sup> ADS = Average Daily Steps: The total number of steps for a patient over a month divided by the number of active days.
<sup>3</sup> ASE = Average Steps per Epoch: The total number of steps in a day divided by the number of active epochs. This daily average is then averaged over all active days in the month. Wearable metrics represent a minimum of 8 epochs (4 hours) worth of data per day.

Table 1 depicts the ambulation metrics aggregated at baseline. The DMD 6MWT and patient data were lower across all three parameters, in keeping with disease-related expectations. Accelerometer and average 6MWT data were similar for the JIA and NP-C groups although the range of 6MWT results was wider for the NP-C cohort.

### Quantitative results

Non-parametric rank tests revealed no significant difference between disease groups for the 6-minute walk test (χ^2^(2) = 4.904, *p* = 0.086). Analysis of the wearable metrics with the Kruskal-Wallis test showed statistically significant differences between the three disease groups for the ADM (χ2(2) = 8.846, *p* = 0.006), ADS (χ2(2) = 7.210, *p* = 0.019) and ASE (χ2(2) = 7.411, *p* = 0.017).

Subsequent analysis with Dunn’s multiple comparison test showed a statistically significant difference in the ADM between the DMD and JIA cohorts (*p* = 0.018). No statistically significant differences were identified between the NP-C/DMD or NP-C/JIA cohorts (*p* = 0.055 and *p* > 0.999 respectively).

In addition, a statistically significant difference for ADS was identified between the NP-C and DMD cohorts (*p* = 0.032). No statistically significant differences were identified between the NP-C/JIA and DMD/JIA cohorts (*p* = 0.891 and *p* = 0.138 respectively).

Lastly for the ASE metric a statistically significant difference was identified between the DMD and JIA cohorts (*p* = 0.0320), but no statistically significant differences were observed between the NP-C/DMD and NP-C/JIA cohorts (*p* = 0.108 and *p* > 0.999 respectively).

The relationship between the 6MWT and wearable metrics was assessed with Spearman’s rank correlation coefficient (α=0.05). In the NP-C, DMD and JIA cohorts there were no statistically significant correlations identified for the ADM and ASE metrics. However, within the JIA cohort, a moderate and statistically significant correlation was identified between the 6MWT and the ADS metric (r_s_=0.68, *p*=0.035).

### Operational feasibility results

As demonstrated in Table 1, the results of the pilot studies show that the collection of data over a month-long period, across three different disease areas, using wearable sensor technology is feasible however data capture was not sustained for months 2-3. Only four patients continued to be highly engaged and long-term users. Patient sensor adherence was limited in all three groups, and particularly significant in the JIA group (Table 2). An overview of disease specific issues with the technology and their respective consequences are presented in Table 3.

**Table 2:** Disease group adherence in days to the wrist worn wearable device per disease, per month.

| Disease group<br>adherence | Month 0-1 | Month 1-2 | Month 2-3 | N= highly engaged<br>patients (> 3<br>months data) |
| --- | --- | --- | --- | --- |
|  | (mean number of days per month [SD]) |  |  |  |
| NP-C | 20.3 (±7.4) | 15.5 (±12.0) | 8.5 (±9.8) | 2 |
| DMD | 13.0 (±9.0) | 5.3 (±8.2) | 7.3 (±9.0) | 2 |
| JIA | 13.7 (±8.8) | 5.5 (±9.8) | 3.3 (±7.3) | 0 |
| Combined | 14.8 (±8.6) | 7.5 (±10.2) | 5.6 (±8.2) | 4 |

**Table 3:**
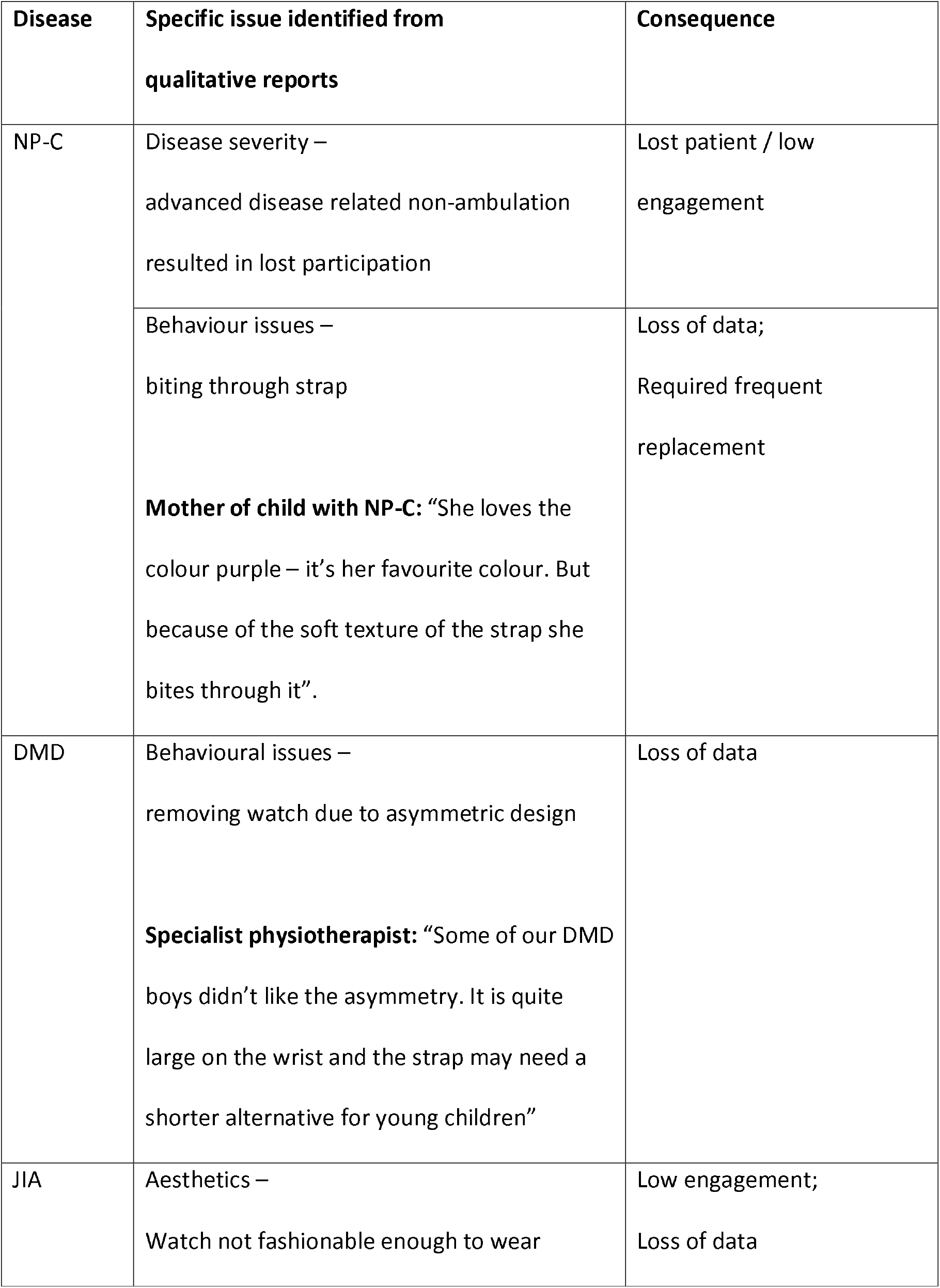
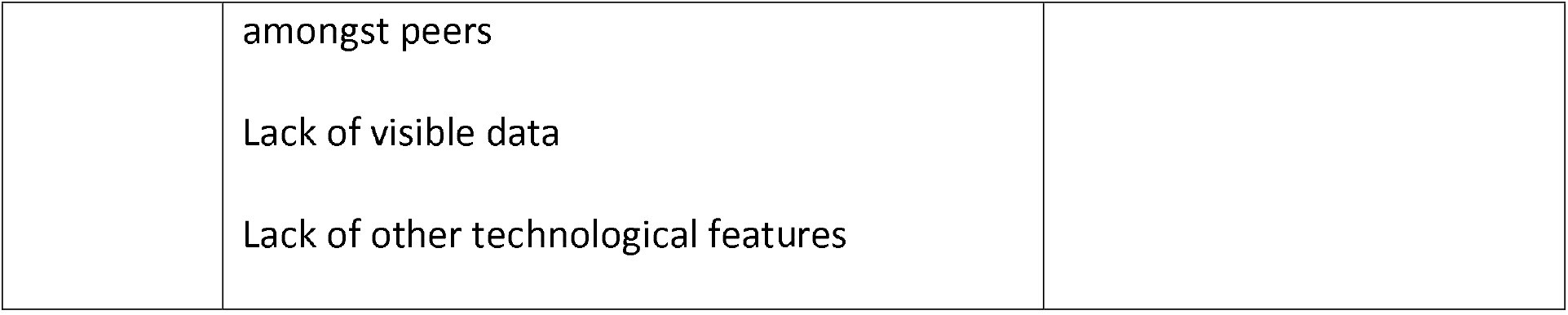
Overview of disease specific issues with the technology and their respective consequences.

## Discussion

Normal childhood activity is key to maintaining childhood health, in particular bone health, muscle development, physical and biopsychosocial health. However, physical activity is a difficult construct to measure during intermittent snapshot clinical assessments. Novel wearable technologies offer a feasible and tolerable opportunity to gather continuous real-world physical activity data in CYP with chronic conditions affecting gait and mobility.

CYP and their families appeared keen to engage with wearable technologies at study recruitment, and setting up the device of the app and device was relatively straightforward. However, patient sensor adherence was limited in all three disease groups; contributing factors included issues relating directly or indirectly to the disease, issues with device function and design and limited patient / family access to the data. Direct patient / family data view was purposefully not included in this deployment to avoid the possibility of families observing functional deterioration in DMD and NP-C or altering activity behaviour. Non-adherence became more apparent later in the study, suggesting that maintaining high levels of compliance might require shorter study durations. Sharing wearable data with CYP and their families in non-degenerative conditions such as JIA is technically possible and may be important in future studies exploring the impact of physical activity interventions. Involving CYP and their families from conception onwards will ensure that future studies are designed to use these technologies to collect comprehensive patient data as effectively as possible.

In addition to the operational feasibility results, this study demonstrates that the acquisition of quantitative wearable sensor data in rare disease populations can add important real-life insights to the snapshot data collected during clinic visits (e.g. 6MWT).

6MWT results are lower than expected in all three cohorts. [20] ADS counts are also lower than expected for healthy age-matched children in all three disease areas. According to Tudor-Locke et al, [21] 6 to 11 year-old boys are expected to average 13,000-15,000 steps per day, 6 to 11-year-old girls 11,000-12,000 steps and 12 to19 year-old adolescents 10,000-12,000 steps, of which 6000 should be in moderate to vigorous physical activity. ADS data presented for all three cohorts participating in this study are consistent with those expected for individuals living with disability and/or chronic illness at 6000-8000.

Impaired physical function is a well-recognised challenge for CYP living with JIA [21]. Although medical management has advanced significantly over the past fifteen years, many CYP with JIA continue to experience episodes of active disease despite intensive treatment schedules [22]. A 2018 North American survey identified impaired ability to walk, run or stand as one of the most impactful symptoms reported by young people with JIA [23]. Identifying, understanding and addressing reduced physical activity levels in children and young people with JIA is therefore a key component of patient-centred clinical care.

In DMD the 6MWT aims to capture functional capacity and endurance, a different concept to activity or aerobic capacity. Other studies have demonstrated a correlation of the 10-metre walk run test and stride length with activity in DMD and also noted decline over time in the population [24] which is in line with natural progression of the disease.

Wearable sensor data measure longitudinal physical activity, providing an overview of function that may not be apparent in snapshot clinical or lab-based settings. In this study, the wearable sensors detected important differences in physical activity between the three disease cohorts, which could not be identified by the 6MWT alone. Capturing longitudinal physical activity data is a key participatory outcome for CYP with chronic illnesses. Employing modern technologies such as wearable activity monitors in future patient-centred clinical trials has the potential to significantly expand our understanding of the lived experience of chronic paediatric illness, potentially highlighting areas for important clinical interventions.

One major limitation of this study is the relatively small number of participants enrolled. Although this is typical in the rare disease space, this led to an even smaller number of patients (particularly NPC patients) collecting wearable data. Whilst it is clear that wearable technologies have great potential in paediatric chronic illness, our numbers are too small to allow meaningful conclusions about activity in our three disease cohorts. This is compounded with the use of 30-minute epochs and the central tendency measures of step counts used in this feasibility study, which collapses over the large volumes of collected data and results in a summary statistic.

A larger study using the more sophisticated technologies that are now available would provide data sets that are more representative of the respective patient populations. These data would provide clinical teams with valuable information about the prevalence and patterns of reduced physical activity in chronic childhood illness, ultimately empowering clinical teams to provide higher quality, holistic clinical care.

## Conclusion

This study demonstrates the feasibility of remote, continuous, long time series monitoring of patients’ physical activity in different paediatric diseases and suggests that wearable data capture can illustrate patterns of physical activity. While further work is required to correlate the wearable device data with other clinical markers and self- or parent-report, there is an increased momentum in the use of wearable sensors and apps as outcome measures. Future studies, involving CYP and their families from conception onwards, have the potential to achieve age-appropriate capture of meaningful and high quality data, adding crucial information to our understanding of the impact of chronic paediatric illness on everyday life.

## Data Availability

All data are available upon reasonable request

## Acknowledgments

The authors would like to acknowledge Duchenne UK for their funding. Niemann Pick Association UK and International Niemann Pick Disease Alliance (INPDA) for their collaboration and Professor Paul Gissen for on-going support.

Acknowledge research nurses Claire Duong at Newcastle.

Newcastle Physiotherapists: Jassi Sodhi, Dionne Moat, Robert Muni Lofra, Meredith James and Michelle McCallum.

## Competing Interests, Funding and all other required statements

Elin Haf Davies and Cecile Ollivier are employees of Aparito and holds shares.

Part of this work was supported by Actelion Pharmaceuticals.

The authors received no funding for the writing of this manuscript.

## Contributors

EHD, AM, VH, FME and AD drafted the initial protocol; CO, EHD and OA drafted the initial paper; AM, VH, FME and AD revised the paper; and all the authors approved the final version

## What is already known on this topic

- Physical activity is pivotal for the healthy development of children.
- Paediatric diseases in which motor function is affected could place patients at risk of developing comorbidities later on in life.
- Wearable technologies have the potential to accurately measure longitudinal ambulation aptitude across paediatric populations.

## What this study adds

- Wearable, digital technology can provide a panoramic assessment of patient ambulatory capacity.
- Wearable, digital technology can potentially serve as an adjunct or alternative to conventional methods employed in clinical trials through delivering real time, remote monitoring.

